# The further development of cold plasma technology: The effectiveness of a contactless, indirect atmospheric cold plasma method for germ reduction on surfaces in vitro and in vivo

**DOI:** 10.1101/2024.10.12.24315382

**Authors:** Ulrich Schmelz, Tom Schaal, Gilbert Hämmerle, Tim Tischendorf

**Affiliations:** University of Fulda, Fulda, Germany; Faculty of Health and Healthcare Sciences, University of Applied Sciences Zwickau, Zwickau, Germany; DGKP, ZWM, AZWM, Head of Wound Outpatient Clinic LKH Bregenz

**Keywords:** Cold plasma, Wound treatment, Chronic wounds, Germ reduction

## Abstract

This study investigates the antimicrobial potential of an indirect cold plasma method for the treatment of wounds. Indirect plasma methods differ from direct methods in that the cold plasma does not come into direct contact with the surface to be treated. The indirect plasma method described here has been implemented in the PLASMOHEAL device. The device generates an aerosol of liquid particles, which is conditioned with plasma reaction products and passed over the areas to be treated without contact. In vitro tests show a significant germ reduction of 3.4 to 4.5 log levels against various microorganisms. In vivo tests on volunteers demonstrate a reduction in E. coli contamination of 4.06 to 5.15 log levels. These results show that indirect plasma methods can achieve equivalent effects to direct methods. The highly effective, pain-free treatment at moderate costs make the indirect plasma method a promising option in modern wound care.

## Introduction

### What is a physical “plasma“?

“Plasma” in the sense of the physical definition, represents the state of a conductive gas. In the plasma state, a gas that is practically non-conductive under normal conditions is partially ionized by an ignition pulse. The gas ions are then electrically charged particles, whereby the gas assumes the state of an ionic conductor (2nd order conductor). This state is maintained by a further supply of electrical energy.

The ionization of gas molecules leads to a transfer of electrical energy to the gas molecules. The ionized gas molecules can transfer this energy, for example to surfaces. As a result, surfaces or even liquids or the surrounding air can be modified [1].

Plasma can occur at atmospheric, elevated or reduced pressure. Plasma can also be altered by the addition of heat energy [2]. Furthermore, plasmas are classified according to the reacting gases; in the process currently under consideration, oxygen and water vapor are brought to react as reactants in the plasma.

The process considered in this paper takes place at room temperature and atmospheric pressure, hence the term ’atmospheric cold plasma’. The gas used for the plasma reactions is ambient air, which contains approximately 21 percent oxygen and water vapour up to the (temperature-dependent) condensation limit. Accordingly, the ambient air supplies the gases (oxygen and water vapor), which are required for the plasma reaction [3]. Plasma reactions can be direct or indirect reactions (Figure 1).

**Figure 1.**
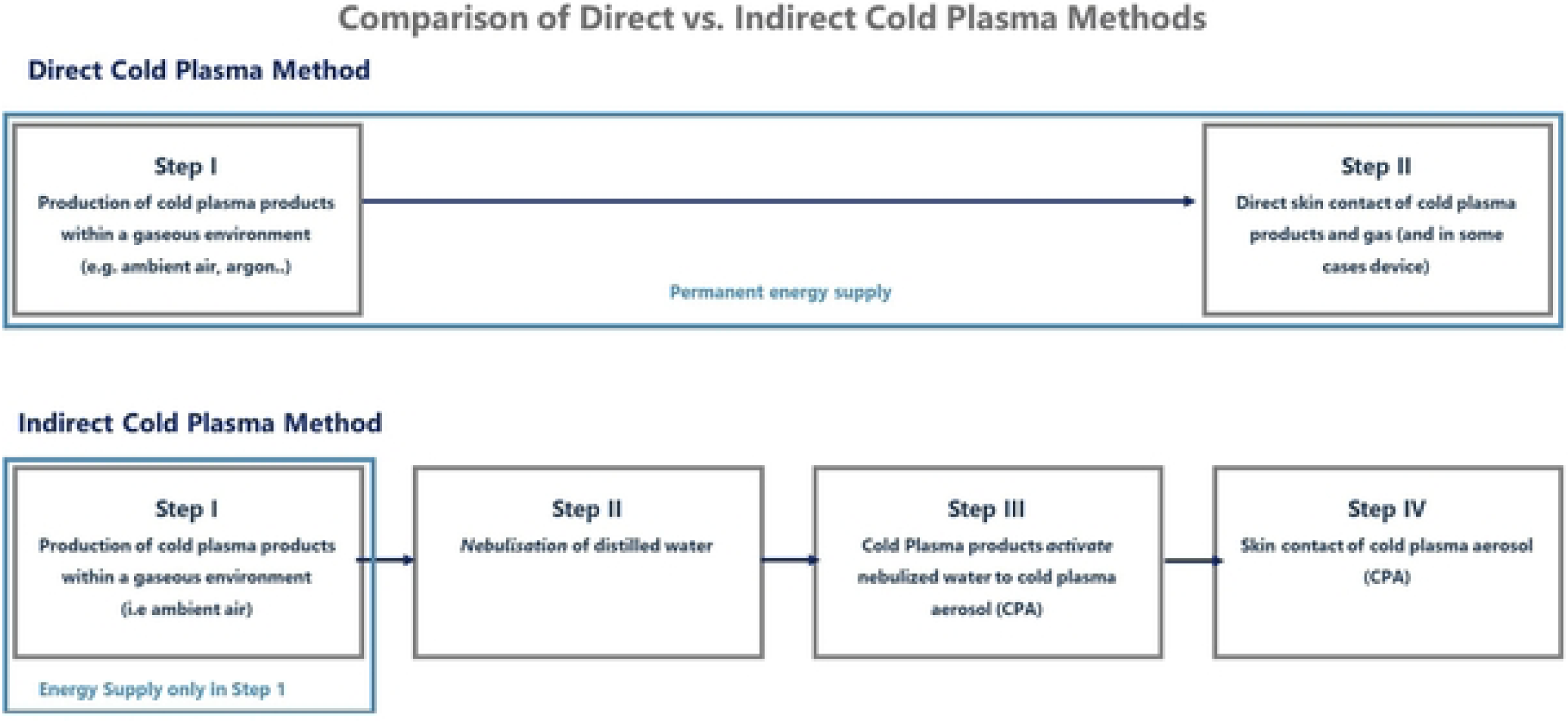
Forms of plasma reactions: direct and indirect plasma reactions.

Until 2024, published studies have primarily documented direct plasma procedures or a combination of direct and indirect procedures in the field of wound sanitation [4]. The antimicrobial effect of indirect cold plasma methods on surfaces was basically shown in 2024 [5]. In a direct plasma reaction, the direct contact of the plasma with a surface leads to a modification of this surface. Energy is only required for the plasma reaction (Figure 1: direct method: step 1 + step 2; indirect method: step 1). Direct methods are operating in consideration of the surface. So direct plasma reactions are in contact to the surface needs to treat.

Additional steps are found with indirect plasma methods. In the case of indirect plasma reactions, additional steps are added that decouple the plasma reaction from the surface to be treated. Due to decoupling, the plasma reaction always takes place at a distance from the surface to be treated. This means that the energy required for the plasma reaction can be outsourced to an external device (Figure 1: step 1; indirect method).

In an indirect plasma reaction, the process takes place outside the relevant surface. Such an indirect plasma method is used, for example, in the “PLASMOHEAL” device (WK-MedTec GmbH, Bückeburg). The device produces the cold plasma reaction products, primarily hydroxyl radicals, which are generated by a targeted flow of ambient air via a corresponding plasma source [3]. These are generated on the basis of the defined potential difference (voltage; here: 1.45 kV as the effective value of the alternating voltage), the frequency (38 kHz) and the frequency form (sinusoidal frequency). Ozone is only present in very small traces.

Then distilled water is nebulized using an ultrasonic module. Ambient air which has been enriched by cold plasma products activate the nebulized water to a cold plasma aerosol (CAP). The plasma reaction products of the air lead to an increase in the electrophysical potential of the nebulized liquid without any material change [2] (Figure 2).

**Figure 2.**
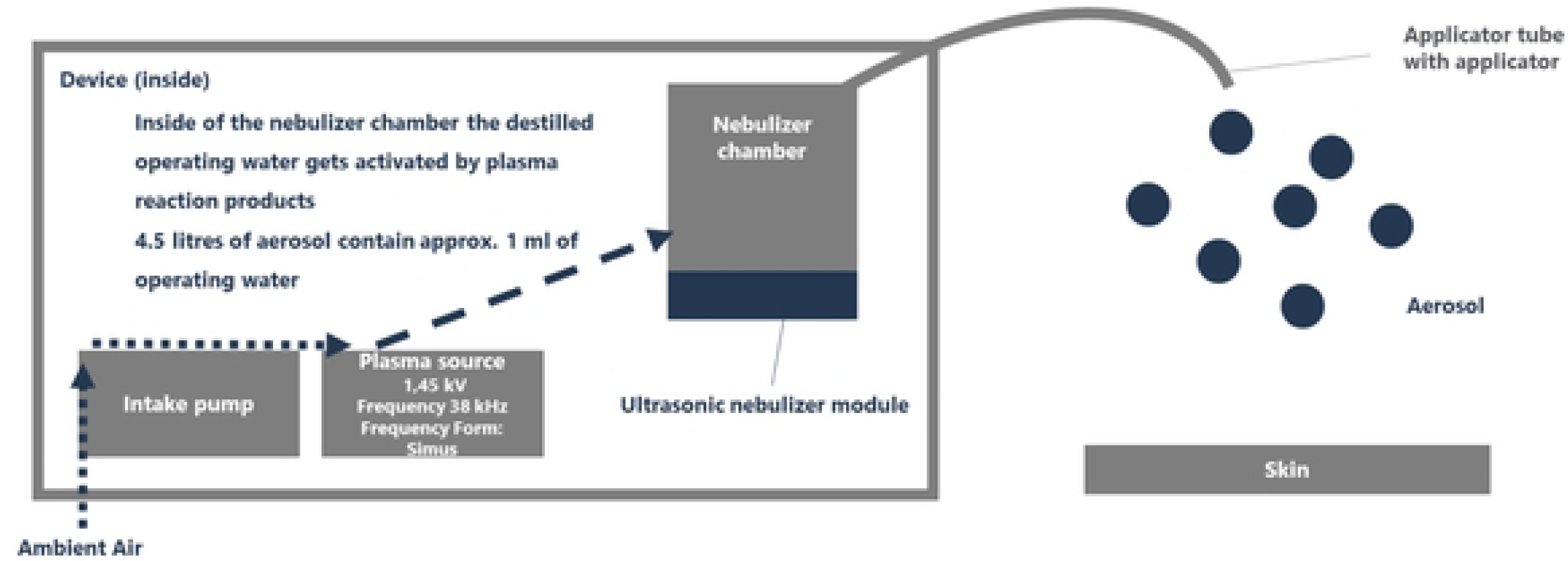
Functional diagram of an indirect cold plasma method using the example of PLASMOHEAL. Note. Distilled water is used as the process water for aerosol formation. 4.5 liters of aerosol contain 1 mL of process water as an aerosol component.

The aerosol is then passed over the areas to be disinfected (here: skin, in particular wounds) for three minutes at a distance of 7.5 cm.

### Effect of direct and indirect plasma methods against microorganisms on surfaces

Both direct and indirect plasma methods are surface-active methods. This focuses on the transient flora, which is preferably found on the surface of the skin, mucous membranes or wounds, while the resident flora (local flora, microbiome) is also localized in deeper layers. The plasma reaction products are effective on the surface of the skin and therefore not in the tissue. In the case of wounds, the surface is interrupted so that the method could also be effective in the contact area of the aerosol in superficial horny layers.

Cold plasma products (contain in aerosols or in direct reaction) support the disruption of the redox processes of microorganisms. Specifically, a “short circuit” of the cell membrane occurs, which leads to a collapse of the resting potential between the extracellular and intracellular space, whereby all transmembrane transport processes in the microorganism are damaged and the microorganism is destroyed [4]. As a result, there is generally no longer any potential for infection. The shown short circuit can be described as a “selective short circuit”, as the human cells are practically not affected (Figure 3).

**Figure 3.**
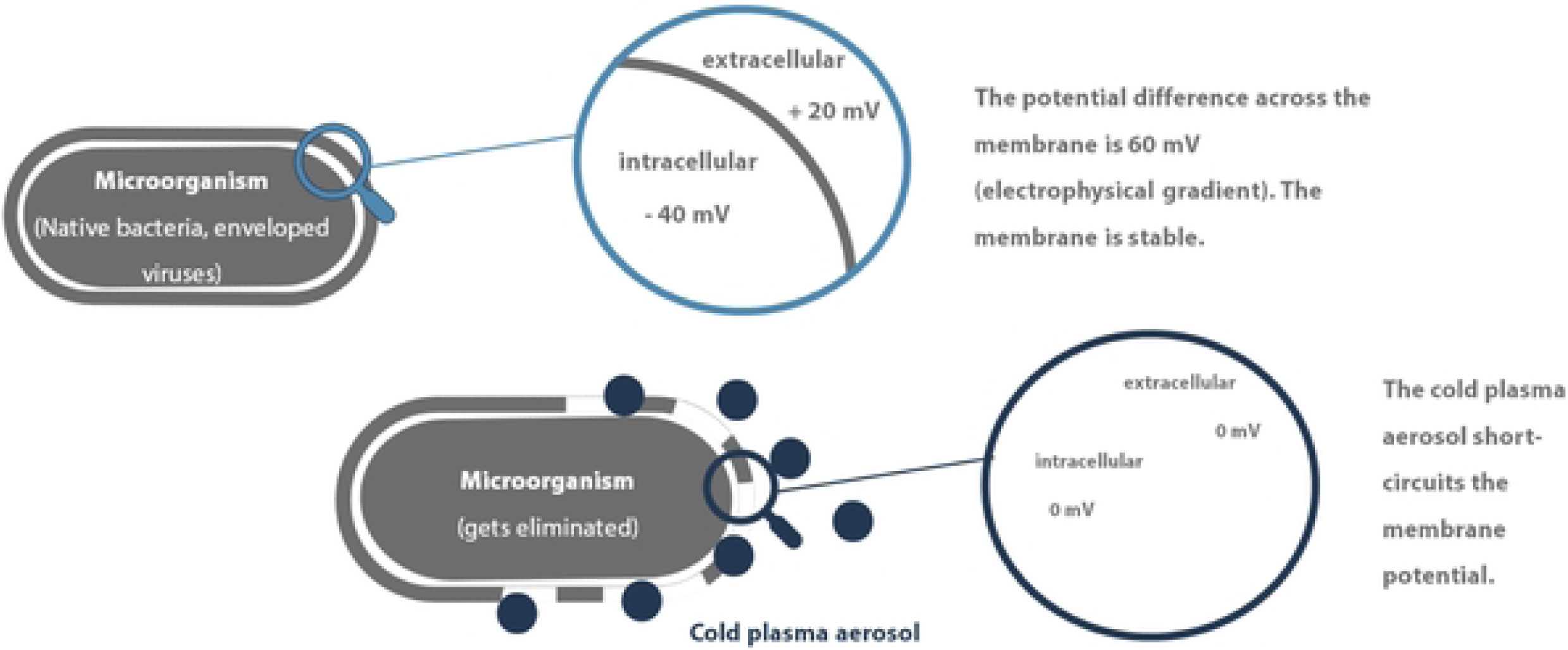
Cold plasma eliminates microorganisms by a short circuit of the membrane potential.

### Absence of toxicological effects on human tissues and cells

The effects occurring in microorganisms are not observed in higher organisms. Toxicologically relevant samples or by-products are not found in indirect plasma processes [6]. To substantiate this, toxicity assessments can provide support. As an example, toxicity tests such as the Ames test (according to EN ISO 10993-3) and a cytotoxicity test (according to EN ISO 10993-5) were performed for the CAP on which the device is based. Both assessments showed the absence of mutagenic effects in human cells. Furthermore, no adverse effects were observed with multiple clinical applications of direct cold plasma systems in wound treatment. The presence of antioxidant enzymes (catalase, superoxide dismutase, alpha-1-antitrypsin) in tissues and organisms that effectively neutralize plasma reaction products before damage to the cell membrane of tissue cells is assumed [6]. It is important to note that there are no recognizable acute or chronic toxic effects on higher organisms (eukaryotic tissues: plants, animals, humans). This lack of effect can be attributed to the presence of beforehand mentioned antioxidant enzymes (catalase, superoxide dismutase, alpha-1-antitrypsin) in these tissues and organisms [6].

Hence, the indirect plasma process appears to be able to eliminate microorganisms without harming humans, thereby reducing the risk of infection, which has already been proven in comparison with direct plasma processes.

### Proof of efficacy of direct plasma methods in the field of wound rehabilitation

A germ-reducing effect of the direct plasma is generally recognized. In studies in vivo (on 18 patients), a reduction of germs in the wound was observed in swabs, regardless of the localization and size of the wound, as well as previous illnesses and cofactors (e.g. tobacco abuse, cardiovascular diseases) [7].

In this study, microbiocidal activity against both Gram-positive cocci bacteria and Gram-negative rod bacteria (both groups include the most common human infectious agents) is sufficiently demonstrated. At the same time, Daeschlein et al. have shown that there are no differences in potency regarding inhibitor-resistant variants of certain microorganisms (e.g. MRSA, MRGN). The microorganism is damaged “in toto” by the electrophysical reaction, which means that a mutation of metabolic pathways has no influence on the microbiocidal potency [7]. Furthermore, Zimmermann et al. describe an in vitro virucidal effect against adenoviruses in the range of approximately 3 log levels [8].

The evaluation of the results of Daeschlein et al. and Zimmermann et al. leads to the conclusion that a microbiocidal effect of > 3 log levels can be expected from the direct plasma methods.

The germ-reducing effect is also indirectly demonstrated by the observation that the pH value of the wound drops when treated with direct cold plasma methods. The typical bacterial wound infection pathogens result in an increase in the pH value of the wound to a range of 8 to 10. The subsequent killing of these alkalizing microorganisms as a result of the cold plasma method then brings about the pH reduction required for optimal wound healing [9].

In addition, a considerable acceleration of wound healing was observed with the use of direct plasmas [10; 11; 12; 13, 14].

According to Strohhal et al. 2022, it is shown that wounds treated with plasma products have a higher probability of healing (approx. 60 per cent higher compared to supportive therapy alone). The individual localization and genesis of the wound is practically irrelevant. Furthermore, application studies with immunological skin diseases (using the example of rosacea) have shown that the method is also forward-looking in the treatment of such immunological diseases [15]. Finally, it should be noted that the claimed effectiveness of the direct methods can be attributed to the antimicrobial effect. The studies of the effectiveness of the direct methods would therefore also be applicable to the indirect methods.

### Derivation of the research question

The antimicrobial potential of the CAP could be used for specialized disinfection, for example of wounds. The cold plasma disinfection methods for wounds used to date - with acceptable results - are almost exclusively direct-acting methods with a direct plasma jet or plasma field. If an equivalent microbiocidal efficacy of indirect plasma methods to direct plasma methods can be demonstrated, it can be concluded that indirect plasma methods also have an equivalent effect in the field of wound decontamination.

This leads to the following research question: Does the indirect cold plasma process have at least an equivalent antimicrobial effect compared to direct cold plasma processes?

To provide this proof, the microbiocidal efficacy of an indirect plasma method is tested in vitro and in vivo in accordance with existing test standards.

## Methods

### Methodology of *in vitro* testing of the microbiocidal effect

An in vitro test of the germ reduction performance against five normatively defined microorganisms (Staph. aureus ATCC 6538, Staph. epidermidis ATCC 14990, E. coli NCTC 19538, Pseudomonas aeruginosa ATCC 10145, Candida albicans ATCC 10321, BS) was carried out in accordance with DIN spec. 91315:2014. The identity of the test organism was checked by using Gram staining in conjunction with transmitted light microscopy and by determining the metabolic performance of the test organism in relation to various substrates. Furthermore, the test organism was identified as indole-forming from tryptophan (indole-positive). The universal medium Caso agar in 60 mm Petri dishes was used as a culture medium for cultivating the inoculates of the samples. After checking their identity, the microorganisms were cultivated as a surface culture and washed with sterile and pyrogen-free NaCl solution 0.9 per cent to obtain a microbial suspension with a turbidity corresponding to McFarland Standard eight. The germ suspensions were then applied to stainless steel test specimens (100*10*1mm) with a roughness depth of 100µm using a sterile swab. The stainless steel test specimens were prepared by immersion for five seconds in the test germ suspension, draining and air drying for two hours. These prepared test specimens were then exposed to the PLASMOHEAL process. The test specimen was exposed to the aerosol of the device for three minutes. The microorganisms were then washed off by resuspending them in 10 mL of sterile and pyrogen-free NaCl solution 0.9 per cent in a sterile test tube by shaking for 30 seconds using a VORTEX 3 shaker. From this suspension (undiluted, dilution factor therefore 10^0^) a geometric dilution series (in powers of ten) up to 10^-3^ is created. A volume of 0.1 mL is taken from each dilution stage and applied to the culture medium suitable for the respective test organism. The volume of 0.1mL is homogeneously distributed on the culture medium using a Drigalski spatula (Figure 4).

**Figure 4.**
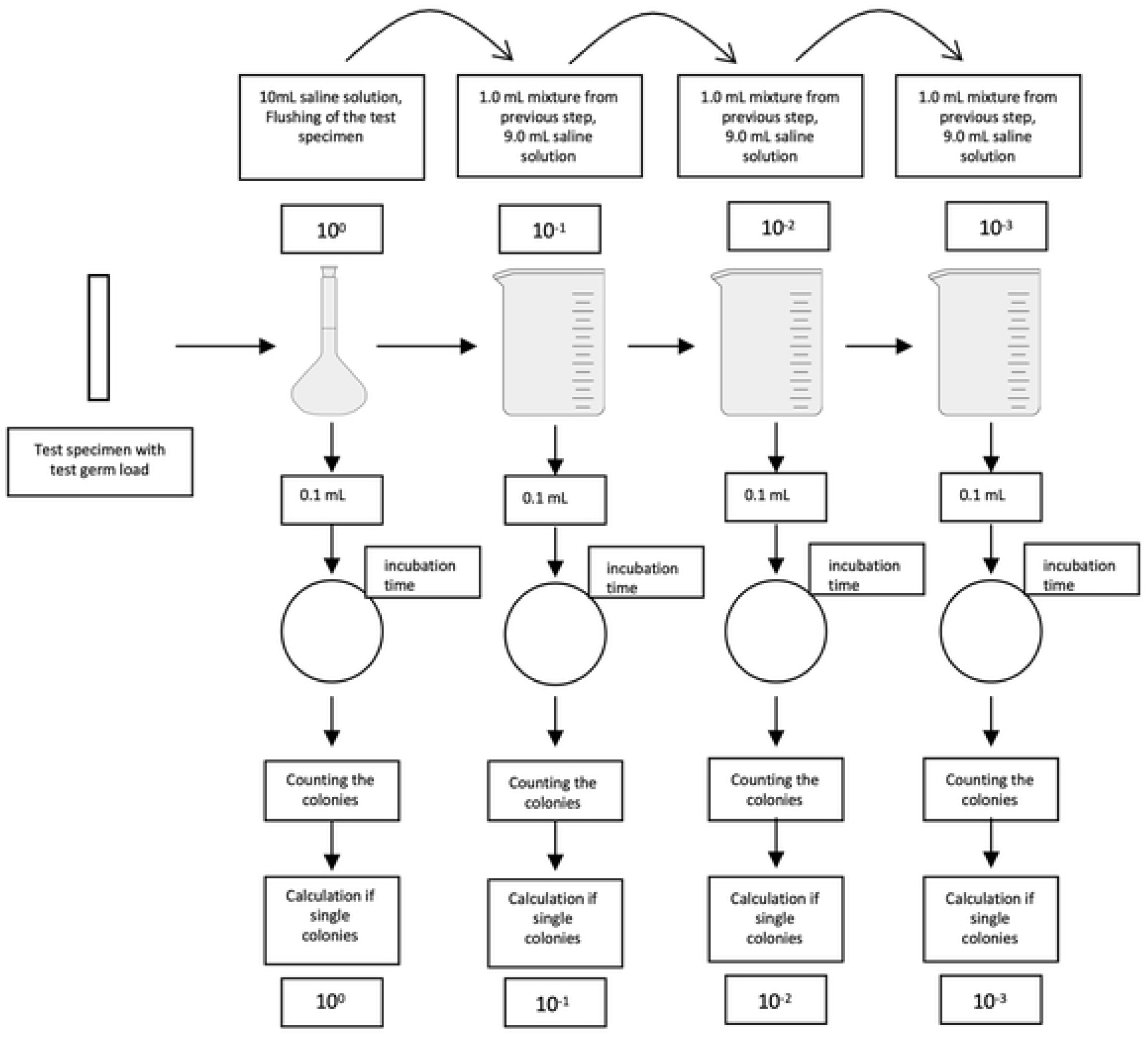
Overview geometric dilution row.

In parallel, an untreated test specimen is also analyzed for each test organism (Table 1).

**Table 1.** Microorganisms used for the in vitro test in accordance with DIN spec. 91315.

| Microorganisms | Germ identification | Culture media with identification number | Incubation time and temperature |
| --- | --- | --- | --- |
| Staphylococcus aureus | ATCC 6538 | Mannitol-salt-agar<br>No.1050 | 48h / 36°C |
| Staphylococcus epidermidis | ATCC 14990 | Caso-agar<br>No. 4020 | 48h / 36°C |
| Escherichia coli | NCTC 19538 | Colichromer-agar<br>No. 4028 | 48h / 36°C |
| Pseudomonas aeruginosa | ATCC 10145 | Cetrimid-agar<br>No. 4025 | 48h / 36°C |
| Candida albicans | ATCC 10321 | Sabouraud-agar 2%<br>No. 4130<sup>1</sup> | 72h / 28°C |

Note. <sup>1</sup>The microbiological culture media for this analysis were obtained from Dr. Möller & Schmelz, Göttingen, Germany.; ATCC = American Type Culture Collection. NCTC = National Collection of Type Cultures.

After the incubation time required for the respective test germ in the incubator, the colonies of the culture medium of the dilution stage containing ten to 100 colonies are counted. Considering the dilution level, the fact that 0.1mL of 10mL per dilution level was applied to the culture medium and the initial volume of 10mL (in which the washout took place), the bacterial count per test specimen is calculated. After forming the decadic logarithm of the bacterial count, the log of the bacterial count after exposure is subtracted from the logarithm of the bacterial count without exposure. This is used to calculate the log reduction factor, which indicates by how many powers of ten an initial bacterial load is reduced by the method under consideration.

### Methodology of *in vivo* testing of the microbiocidal effect

Based on the results of the in vitro test, the germ reduction performance was tested in the in vivo test in accordance with EN 1500 using the test germ Escherichia coli K12. EN 1500 is used for microbiocidal testing of hand sanitizers and therefore allows in vivo testing on test subjects. The EN 1500 standard on which the test is based describes the wetting of the fingertips as the location of the wash-off, because these can be mobilised mechanically by rubbing with the opposing thumb. The hands were immersed in the test germ suspension for five seconds in the area of the fingers, then drained and loaded with the test germ for three minutes. Before and after application of the PLASMOHEAL method, the bacterial count of the skin of the hands was methodically determined in accordance with EN 1500, with both hands contaminated by the bacterial suspension in the area of the fingers. The “pre-value” without the use of the device was determined from the left hand, then the right hand was exposed to the aerosol in the area of the contaminated fingers for three minutes. The “after value” was then determined from the exposed right hand. The dilution series and cultivation were carried out analogue to the method described above for the in vitro test of the microbiocidal effect. After cultivation and evaluation, the log reduction factor of the cold plasma process when applied to human skin was determined. For evaluation, the test result of the PLASMOHEAL method was compared with the log reduction factor of standard alcohol as a reference method following the analogy of EN 1500.

EN 1500 specifies E. coli K12 as the test germ, as this microorganism is apathogenic (it is also a natural commensal of the human colon, among other things) and furthermore its tenacity (resistance to inactivation) includes sufficient obligate pathogenic germs, E. coli is a sufficient disinfection surrogate and at the same time there is no infectiological risk. E. coli on the hand is therefore used as a surrogate for a contaminated wound. Therefore, the performance of the disinfection efficacy test is not a clinical test in the sense of requiring approval by an ethics committee. Rather, this test is a standard procedure, i.e. “state of the art or science”, which is required according to EN 1500 in order to determine the aforementioned disinfection efficacy in a technically reproducible manner. The test is carried out in this form many times a day in Germany.

The test subjects have declared to the person carrying out the test that the test is voluntary, furthermore there are no direct dependencies, and the test subjects also declared that they were not, did not want to be, or could not be pregnant. We confirm that all methods of this study were conducted in accordance with the relevant guidelines and regulations. The participants have given their informed consent to participate in the study in advance. Written informed consent to participate was obtained from the participants. The consents were documented in a standardized laboratory list and signed by the participants. No minors were included. A written exemption from ethics approval was obtained from the institutional review board, as the test was carried out in accordance with the requirements of the EN 1500 test standard and its effectiveness was confirmed in laboratory tests.

## Results

The aim of this study is to investigate the germ-reducing effect of the PLASMOHEAL process both in vitro and in vivo. By analyzing in detail the results of the in vitro tests on the germ-reducing effect of the PLASMOHEAL procedure (Table 2) and the in vivo tests (Table 3), the dynamic effect of this innovative wound treatment method is comprehensively examined. The germ-reducing properties of the procedure are presented in detail at various levels of testing.

**Table 2.** Results of the in vitro test of the germ-reducing effect of the PLASMOHEAL process.

| No. | V1 | V2 | CfU/ Agar plate | CfU/ Approach | Dilution | CfU/ Proband | Log CfU/ Proband | Log Reductions faktor |
| --- | --- | --- | --- | --- | --- | --- | --- | --- |
| **Staphylococcus aureus 3min:** |  |  |  |  |  |  |  |  |
| 1.1.A-3 | 10,0 mL | 0,1 mL | 87 | 8,700 | 10,000 | 87,000,000 | **7.94** |  |
| 1.1.B-3 | 10,0 mL | 0,1 mL | 28 | 2,800 | 1 | 2,800 | **3.45** | **4.49** |
| 1.2.A-3 | 10,0 mL | 0,1 mL | 56 | 5,600 | 10,000 | 56,000,000 | **7.75** |  |
| 1.2.B-3 | 10,0 mL | 0,1 mL | 26 | 2,600 | 1 | 2,600 | **3.41** | **4.33** |
| 1.3.A-3 | 10,0 mL | 0,1 mL | 78 | 7,800 | 10,000 | 78,000,000 | **7.89** |  |
| 1.3.B-3 | 10,0 mL | 0,1 mL | 22 | 2,200 | 1 | 2,200 | **3.34** | **4.55** |
| **Staphylococcus epidermidis 3min:** |  |  |  |  |  |  |  |  |
| 1.1.A-3 | 10,0 mL | 0,1 mL | 56 | 5,600 | 10,000 | 56,000,000 | **7.75** |  |
| 1.1.B-3 | 10,0 mL | 0,1 mL | 31 | 3,100 | 1 | 3100 | **3.49** | **4.26** |
| 1.2.A-3 | 10,0 mL | 0,1 mL | 44 | 4,400 | 10,000 | 44,000,000 | **7.64** |  |
| 1.2.B-3 | 10,0 mL | 0,1 mL | 25 | 2,500 | 1 | 2,500 | **3.40** | **4.25** |
| 1.3.A-3 | 10,0 mL | 0,1 mL | 76 | 7,600 | 10,000 | 76,000,000 | **7.88** |  |
| 1.3.B-3 | 10,0 mL | 0,1 mL | 37 | 3,700 | 1 | 3,700 | **3.57** | **4.31** |
| **Escherichia coli 3min:** |  |  |  |  |  |  |  |  |
| 1.1.A-3 | 10,0 mL | 0,1 mL | 92 | 9,200 | 10,000 | 92,000,000 | **7.96** |  |
| 1.1.B-3 | 10,0 mL | 0,1 mL | 54 | 5,400 | 1 | 5,400 | **3.73** | **4.23** |
| 1.2.A-3 | 10,0 mL | 0,1 mL | 78 | 7,800 | 10,000 | 78,000,000 | **7.89** |  |
| 1.2.B-3 | 10,0 mL | 0,1 mL | 34 | 3,400 | 1 | 3,400 | **3.53** | **4.36** |
| 1.3.A-3 | 10,0 mL | 0,1 mL | 65 | 6,500 | 10,000 | 65,000,000 | **7.81** |  |
| 1.3.B-3 | 10,0 mL | 0,1 mL | 39 | 3,900 | 1 | 3,900 | **3.59** | **4.22** |
| **Pseudomonas aeruginosa 3min:** |  |  |  |  |  |  |  |  |
| 1.1.A-3 | 10,0 mL | 0,1 mL | 72 | 7,200 | 10,000 | 72,000,000 | **7.86** |  |
| 1.1.B-3 | 10,0 mL | 0,1 mL | 26 | 2,600 | 1 | 2,600 | **3.41** | **4.44** |
| 1.2.A-3 | 10,0 mL | 0,1 mL | 61 | 6,100 | 10,000 | 61,000,000 | **7.79** |  |
| 1.2.B-3 | 10,0 mL | 0,1 mL | 34 | 3,400 | 1 | 3,400 | **3.53** | **4.25** |
| 1.3.A-3 | 10,0 mL | 0,1 mL | 93 | 9,300 | 10,000 | 93,000,000 | **7.97** |  |
| 1.3.B-3 | 10,0 mL | 0,1 mL | 45 | 4,500 | 1 | 4,500 | **3.65** | **4.32** |
| **Candida albicans 3min:** |  |  |  |  |  |  |  |  |
| 1.1.A-3 | 10,0 mL | 0,1 mL | 78 | 7,800 | 10,000 | 7,800,000 | **6.89** |  |
| 1.1.B-3 | 10,0 mL | 0,1 mL | 31 | 3,100 | 1 | 3,100 | **3.49** | **3.40** |
| 1.2.A-3 | 10,0 mL | 0,1 mL | 59 | 5,900 | 10,000 | 59,000,000 | **7.77** |  |
| 1.2.B-3 | 10,0 mL | 0,1 mL | 27 | 2,700 | 1 | 2,700 | **3.43** | **4.34** |
| 1.3.A-3 | 10,0 mL | 0,1 mL | 86 | 8,600 | 10,000 | 86,000,000 | **7.93** |  |
| 1.3.B-3 | 10,0 mL | 0,1 mL | 29 | 2,900 | 1 | 2,900 | **3.46** | **4.47** |

Note. No. = Identification of the test run, where the first two digits stand for the test person number and 'A' = test run before disinfection (initial bacterial count), 'B' = test run after disinfection (initial bacterial count); V1 = Total volume Approach to leaching; V2 = Volume of the batch from V1, which was applied to the agar plate; CfU = Colony forming Unit.

**Table 3.** Results of the in vivo test of the germ-reducing effect of the PLASMOHEAL process.

| No. | V1 | V2 | CfU/Agar<br>plate | CfU/<br>Approach | Dilution | CfU/<br>Proband | Log<br>CfU/<br>Proband | Log<br>Reductions<br>factor | Mean<br>Values<br>Log RF |
| --- | --- | --- | --- | --- | --- | --- | --- | --- | --- |
| 01 before | 10.0<br>mL | 0.1<br>mL | 46 | 4,600 | 10,000 | 46,000,000 | **7.66** |  |  |
| 01 after | 10.0<br>mL | 0.1<br>mL | 3 | 300 | 1 | 300 | **2.48** | **5.19** | **5.19** |
| 02 before | 10.0<br>mL | 0.1<br>mL | 18 | 1,800 | 10,000 | 18,000,000 | **7.26** |  |  |
| 02 after | 10.0<br>mL | 0.1<br>mL | 5 | 500 | 1 | 500 | **2.70** | **4.56** | **4.56** |
| 03 before | 10.0<br>mL | 0.1<br>mL | 16 | 1,600 | 100,000 | 160,000,000 | **8.20** |  |  |
| 03 after | 10.0<br>mL | 0.1<br>mL | 14 | 1,400 | 1 | 1400 | **3.15** | **5.06** | **5.06** |
| 04 before | 10.0<br>mL | 0.1<br>mL | 23 | 2,300 | 1,000 | 2,300,000 | **6.36** |  |  |
| 04 after | 10.0<br>mL | 0.1<br>mL | 2 | 200 | 1 | 200 | **2.30** | **4.06** | **4.06** |
| 05 before | 10.0<br>mL | 0.1<br>mL | 84 | 8,400 | 10,000 | 84,000,000 | **7.92** |  |  |
| 05 after | 10.0<br>mL | 0.1<br>mL | 5 | 500 | 1 | 500 | **2.70** | **5.23** | **5.23** |
| 06 before | 10.0<br>mL | 0.1<br>mL | 15 | 1,500 | 10,000 | 15,000,000 | **7.18** |  |  |
| 06 after | 10.0<br>mL | 0.1<br>mL | 3 | 300 | 1 | 300 | **2.48** | **4.70** | **4.70** |
| 07 before | 10.0<br>mL | 0.1<br>mL | 4 | 400 | 10,000 | 400,000 | **6.60** |  |  |
| 07 after | 10.0<br>mL | 0.1<br>mL | 1 | 100 | 1 | 100 | **2.00** | **4.60** | **4.60** |
| 08 before | 10.0<br>mL | 0.1<br>mL | 92 | 9,200 | 10,000 | 92,000,000 | **7.96** |  |  |
| 08 after | 10.0<br>mL | 0.1<br>mL | 17 | 1,700 | 1 | 1700 | **3.23** | **4.73** | **4.73** |
| 09 before | 10.0<br>mL | 0.1<br>mL | 39 | 3,900 | 10,000 | 39,000,000 | **7.59** |  |  |
| 09 after | 10.0<br>mL | 0.1<br>mL | 5 | 500 | 1 | 500 | **2.70** | **4.89** | **4.89** |
| 10 before | 10.0<br>mL | 0.1<br>mL | 24 | 2,400 | 10,000 | 24,000,000 | **7.38** |  |  |
| 10 after | 10.0<br>mL | 0.1<br>mL | 1 | 100 | 1 | 100 | **2.00** | **5.38** | **5.38** |
| 11 before | 10.0<br>mL | 0.1<br>mL | 17 | 1,700 | 10,000 | 17,000,000 | **7.23** |  |  |
| 11 after | 10.0<br>mL | 0.1<br>mL | 5 | 500 | 1 | 500 | **2.70** | **4.53** | **4.53** |
| 12 before | 10.0<br>mL | 0.1<br>mL | 11 | 1,100 | 10,000 | 11,000,000 | **7.04** |  |  |
| 12 after | 10.0<br>mL | 0.1<br>mL | 1 | 100 | 1 | 100 | **2.00** | **5.04** | **5.04** |
| 13 before | 10.0<br>mL | 0.1<br>mL | 26 | 2,600 | 10,000 | 26,000,000 | **7.41** |  |  |
| 13 after | 10.0<br>mL | 0.1<br>mL | 2 | 200 | 1 | 200 | **2.30** | **5.11** | **5.11** |
| 14 before | 10.0<br>mL | 0.1<br>mL | 14 | 1,400 | 10,000 | 14,000,000 | **7.15** |  |  |
| 14 after | 10.0<br>mL | 0.1<br>mL | 4 | 400 | 1 | 400 | **2.60** | **4.54** | **4.54** |
| 15 before | 10.0<br>mL | 0.1<br>mL | 14 | 1,400 | 10,000 | 14,000,000 | **7.15** |  |  |
| 15 after | 10.0<br>mL | 0.1<br>mL | 1 | 100 | 1 | 100 | **2.00** | **5.15** | **5.15** |
| 16 before | 10.0<br>mL | 0.1<br>mL | 39 | 3,900 | 10,000 | 39,000,000 | **7.59** |  |  |
| 16 after | 10.0<br>mL | 0.1<br>mL | 9 | 900 | 1 | 900 | **2.95** | **4.64** | **4.64** |
| 17 before | 10.0<br>mL | 0.1<br>mL | 21 | 2,100 | 10,000 | 21,000,000 | **7.32** |  |  |
| 17 after | 10.0<br>mL | 0.1<br>mL | 3 | 300 | 1 | 300 | **2.48** | **4.85** | **4.85** |
| 18 before | 10.0<br>mL | 0.1<br>mL | 14 | 1,400 | 10,000 | 14,000,000 | **7.15** |  |  |
| 18 after | 10.0<br>mL | 0.1<br>mL | 4 | 400 | 1 | 400 | **2.60** | **4.54** | **4.54** |
|  |  |  |  |  |  |  | **Mean**<br>**value**<br>**log RF** | **4.82** |  |
|  |  |  |  |  |  |  | **Median**<br>**log RF** | **4.79** |  |
| **Comparison control with standard alcohol according to EN 1500 (60% n-propanol, 40% water)** |  |  |  |  |  |  |  |  |  |
| 19 before | 10.0<br>mL | 0.1<br>mL | 9 | 900 | 10,000 | 9,000,000 | **6.95** |  |  |
| 19 after | 10.0<br>mL | 0.1<br>mL | 1 | 100 | 1 | 100 | **2.00** |  |  |
| 19 after | 10.0<br>mL | 0.1<br>mL | 3 | 300 | 1 | 300 | **2.48** |  |  |
| 19 after | 10.0<br>mL | 0.1<br>mL | 1 | 100 | 1 | 100 | **2.00** |  |  |
| 19 after | 10.0<br>mL | 0.1<br>mL | 4 | 400 | 1 | 400 | **2.60** |  |  |
| 19 after | 10.0<br>mL | 0.1<br>mL | 2 | 200 | 1 | 200 | **2.30** | **2.28** | **4.68** |

Note. No. = Identification of the test run; V1 = Total volume Approach to leaching; V2 = Volume of the batch from V1, which was applied to the agar plate; CfU = Colony forming Unit; RF = Reduction factor.

## Discussion

The results obtained show a microbiocidal reduction performance of the PLASMOHEAL process tested in vitro against the test germs Staph. aureus, Staph. epidermidis, E. coli, Pseudomonas aeruginosa and Candida albicans of 3.4 to 4.5 log levels. When considering the reduction factors of all five microorganisms tested, a mean value of 4.28 and a median of 4.32 log levels were calculated.

The tested method thus achieves a germ reduction of > 4 powers of ten, which corresponds to a reduction in the germ count density of > 99.99 percent.

The in vivo test with E. coli in accordance with EN 1500 was carried out on the real skin of the test subjects. Due to the similarity in bacterial colonization between skin and wounds, the skin can be regarded as a surrogate for wound testing [16]. A reduction of the initial E. coli contamination in the range between 4.06 and 5.15 log levels was determined. The mean value of log reduction factor across the results of the 18 test subjects was 4.82 log levels, while the median of this group of values was determined to be 4.79 log levels.

The targeted reduction of microorganisms is an important method of disinfection. Disinfection describes an antiseptic process that leads to a state of asepsis and is characterized by the fact that the risk of infection is largely eliminated. This is achieved by reducing the density of microorganisms, which is so low after disinfection that an infection is unlikely [17].

The German Association for Applied Hygiene (VAH) initially recommends the highest possible germ reduction. At least 3 log levels are considered necessary as a quantitative value for germ reducing processes. Depending on the measure and the purpose of use, 4 to 5 log levels are required [18].

The PLASMOHEAL process shows a reduction of 4.28 log levels against the test germs Staph. aureus, Staph. epidermidis, E. coli, Pseudomonas aeruginosa and Candida albicans when tested according to DIN spec. 91315. The in vitro test showed a mean value of 4.28 log levels of germ count reduction. This demonstrates that the disinfection effect of the PLASMOHEAL process is safe under test conditions. Furthermore, the in vivo test based on EN 1500 shows an average reduction performance of 4.82 log levels. This meets both the VAH recommendation of 3 to 5 log levels and the standard alcohol also analyzed as a reference in accordance with EN 1500, which showed a reduction of 4.68 log levels in the current study.

The initially formulated research question can be answered to the effect that the sufficient germ reduction performance of indirect plasma methods is achieved and that indirect plasma methods in the field of wound decontamination can be regarded as at least equivalent to the effect of direct plasma methods. The claimed effect of direct cold plasma methods is the antimicrobial effect. The use of PLASMOHEAL as a proxy for an indirect method shows the equivalence in the efficacy of indirect methods in relation to the assumed efficacy claim of direct methods. Consequently, the studies of direct cold plasma methods can also be transferred to indirect cold plasma methods as part of a metrological traceability.

The energy in the indirect method is only applied in step 1 (Figure 1). As a result, the treatment is painless and non-irritating [9]. By adding the aerosol, a large surface area (up to 10 * 10 cm) can be treated simultaneously within three minutes using the indirect method compared to the direct method.

The overall view of the results and the additional aspects of wound treatment with indirect plasma methods shows that the tested indirect plasma method can be used in a forward-looking manner in wound treatment, particularly in the case of microbiological colonization. The purpose of the analysis was to determine the microbiocidal effect and its quantification, therefore the analysis focused on these parameters. The consideration of other and secondary parameters is another aspect for future analysis.

## Data Availability

All relevant data are within the manuscript and its Supporting Information files.

## Authors’ Contributions

US: Writing – original draft; Formal Analysis; Investigation; Project administration; Conceptualization

TS: Writing – original draft; Formal Analysis; Writing – review & editing

GH: Writing – original draft; Writing – review & editing

TT: Writing – original draft; Visualization; Data curation

## References

[1] Ehlbeck J, Schnabel U, Polák M, Winter J, Von Woedtke T, Brandenburg R, et al. Low temperature atmospheric pressure plasma sources for microbial decontamination. J Phys D Appl Phys. 2011;44(1):013002. 10.1088/0022-3727/44/1/013002.

[2] Lerouge S, Wertheimer MR, Yahia L. Plasma Sterilisation: A Review of Parameters, Mechanisms, and Limitations. Plasmas Polym. 2001;6:175–88. 10.1023/A:1013196629791.

[3] Krömker, W, Schorling, T. Vorrichtung zur Ioniserung von Umgebungsluft [Device for ionising ambient air] (EP 4 175 082 A1). European Patent Office 2023.

[4] Braný D, Dvorská D, Halašová E, Škovierová H. Cold Atmospheric Plasma: A Powerful Tool for Modern Medicine. Int J Mol Sci. 2020;21(8):2932. 10.3390/ijms21082932.

[5] Tischendorf T, Schaal T, Schmelz U. Study on hand disinfection in inpatient geriatric care on the superiority of cold plasma aerosol versus alcohol-based disinfection methods in a parallel group design. Sci Rep. 2024 Sep 17;14(1):21703. doi: 10.1038/s41598-024-72524-7. PMID: 39289454; PMCID: PMC11408535.

[6] Masur K, Schmidt J, Stürmer E, von Woedtke T. Kaltes Plasma zur Heilung chronischer Wunden [Cold plasmas for the healing of chronic wounds]. WundManagement. 2018;12:253–9.

[7] Daeschlein G, Napp M, Lutze S, Arnold A, Von Podewils S, Guembel D, et al. Skin and wound decontamination of multidrug-resistant bacteria by cold atmospheric plasma coagulation. J Ger Dermatol Soc. 2015;13(2):143–9. 10.1111/ddg.12559.

[8] Zimmermann J, Dumler K, Shimizu T, Morfill GE, Wolf A, Boxhammer V, et al. Effects of cold atmospheric plasmas on adenoviruses in solution. J Phys D Appl Phys. 2011;44(50):505201. 10.1088/0022-3727/44/50/505201.

[9] Hämmerle G, Ascher S, Gebhardt L. Positive effects of cold atmospheric plasma on pH in wounds: a pilot study. J Wound Care. 2023;32(9):530–6. 10.12968/jowc.2023.32.9.530.

[10] Rached NA, Kley S, Storck M, Meyer T, Stücker M. Cold Plasma Therapy in Chronic Wounds-A Multicenter, Randomised Controlled Clinical Trial (Plasma on Chronic Wounds for Epidermal Regeneration Study): Preliminary Results. J Clin Med. 2023;12(15):5121. 10.3390/jcm12155121.

[11] Strohal R, Dietrich S, Mittlböck M, Hämmerle G. Chronic wounds treated with cold atmospheric plasmajet versus best practice wound dressings: a multicenter, randomized, non-inferiority trial. Sci Rep. 2022;12(1). 10.1038/s41598-022-07333-x.

[12] Stratmann B, Costea TC, Nolte C, Hiller J, Schmidt J, Reindel J, et al. Effect of Cold Atmospheric Plasma Therapy vs Standard Therapy Placebo on Wound Healing in Patients With Diabetic Foot Ulcers: A Randomized Clinical Trial. JAMA Netw Open. 2020;3(7). 10.1001/jamanetworkopen.2020.10411.

[13] Isbary G, Heinlin J, Shimizu T, Zimmermann J, Morfill G, Schmidt H, et al. Successful and safe use of 2 min cold atmospheric argon plasma in chronic wounds: results of a randomised controlled trial. Br J Dermatol. 2012;167(2):404–10. 10.1111/j.1365-2133.2012.10923.x.

[14] Schachl J, Königshofer M, Stoiber M, Socha M, Grasl C, Abart T, Michel-Behnke I, Wiedemann D, Riebandt J, Zimpfer D, Schlöglhofer T. Cold atmospheric plasma therapy as a novel treatment for Berlin Heart EXCOR pediatric cannula infections. Artif Organs. 2024. doi: 10.1111/aor.14869.

[15] Hofmeyer S, Weber F, Gerds S, Emmert S, Thiem A. A Prospective Randomised Controlled Pilot Study to Assess the Response and Tolerability of Cold Atmospheric Plasma for Rosacea. Skin Pharmacol Physiol. 2023;36(4):205–13. 10.1159/000533190.

[16] Tomic-Canic M, Burgess JL, O’Neill KE, et al. Skin Microbiota and its Interplay with Wound Healing. Am J Clin Dermatol. 2020;21(Suppl 1):36–43. 10.1007/s40257-020-00536-w.

[17] Dunkelberg H, Wedekind S. Eine neue Methode zur Wirksamkeitsprüfung von Sterilgutverpackungen in der Praxis [A new method of testing the effectiveness of sterile packaging in general practice]. Biomed Tech (Berl). 2002;47(11):290–3.

[18] Disinfectant Commission of the VAH (ed.). Anforderungen und Methoden zur VAH-Zertifizierung chemischer Desinfektionsverfahren [Requirements and methods for VAH certification of chemical disinfection procedures]. Chapters 10-11. Available from: https://vah-online.de/files/download/ebooks/eBook_VAH_Methoden_Anforderungen.pdf. Accessed 2024 Aug 17.

